# Risk and protective factors of SARS-CoV-2 infection – Meta-regression of data from worldwide nations

**DOI:** 10.1101/2020.06.06.20124016

**Authors:** Hisato Takagi, Toshiki Kuno, Yujiro Yokoyama, Hiroki Ueyama, Takuya Matsushiro, Yosuke Hari, Tomo Ando

## Abstract

Although it has been reported that coexistent chronic diseases are strongly associated with COVID-19 severity, investigations of predictors for SARS-CoV-2 infection itself have been seldom performed. To screen potential risk and protective factors for SARS-CoV-2 infection, meta-regression of data from worldwide nations were herein conducted. We extracted total confirmed COVID-19 cases in worldwide 180 nations (May 31, 2020), nation total population, population ages 0-14/≥65, GDP/GNI per capita, PPP, life expectancy at birth, medical-doctor and nursing/midwifery-personnel density, hypertension/obesity/diabetes prevalence, annual PM2.5 concentrations, daily ultraviolet radiation, population using safely-managed drinking-water/sanitation services and hand-washing facility with soap/water, inbound tourism, and bachelor’s or equivalent (ISCED 6). Restricted maximum-likelihood meta-regression in the random-effects model was performed using Comprehensive Meta-Analysis version 3. To adjust for other covariates, we conducted the hierarchical multivariate models. A slope (coefficient) of the meta-regression line for the COVID-19 prevalence was significantly negative for population ages 0-14 (–0.0636; *P* = .0021) and positive for obesity prevalence (0.0411; *P* = .0099) and annual PM2.5 concentrations in urban areas (0.0158; *P* = .0454), which would indicate that the COVID-19 prevalence decreases significantly as children increase and that the COVID-19 prevalence increases significantly as the obese and PM2.5 increase. In conclusion, children (negatively) and obesity/PM2.5 (positively) may be independently associated with SARS-CoV-2 infection.

## Introduction

It has been reported that coexistent chronic diseases are strongly associated with coronavirus disease 2019 (COVID-19) severity.^1^ Investigations of predictors for severe acute respiratory syndrome coronavirus 2 (SARS-CoV-2) infection itself, however, have been seldom performed. Although meta-regression has been traditionally utilized to investigate heterogeneity in meta-analysis,^2^ that considering a nation as a study in meta-analysis may be of use to screen potential risk and protective factors for SARS-CoV-2 infection. To screen the factors, meta-regression of data from worldwide nations were herein conducted.

## Methods

We extracted 1) total confirmed COVID-19 cases in worldwide 180 nations on May 31, 2020 from the “COVID-2019 situation reports” (https://www.who.int/emergencies/diseases/novel-coronavirus-2019/situation-reports/) of the World Health Organization (WHO); 2) national total population (thousands) in 2018, life expectancy at birth (years) in 2016, medical-doctor and nursing/midwifery-personnel density (/10,000 population) in 2010–2018, hypertension prevalence (%) in 2015, obesity prevalence (%) in 2016, annual particulate matter 2.5 (PM2.5) concentrations in urban areas (μg/m^3^) in 2016, population using safely-managed drinking-water/sanitation services and hand-washing facility with soap/water (%) in 2017, and daily ambient ultraviolet radiation (J/m^2^) in 1997–2003 from the “World Health Statistics 2020: Monitoring health for the Sustainable Development Goals” (https://www.who.int/gho/publications/world_health_statistics/2020/en/) and “Public health and environment” (https://apps.who.int/gho/data/node.main.122?lang=en) of the WHO; 3) population ages 0-14 and ≥65 (%) in 2018, GDP [Gross Domestic Product] and GNI [Gross National Income] per capita, PPP [Purchasing Power Parity] (current international $) in 2016–2018 (mostly 2018), and diabetes prevalence (%) in 2019 from the World Bank (https://data.worldbank.org/indicator/); 4) inbound tourism (thousands) in 2014–2018 from the UNWTO [World Tourism Organization of the United Nations] (https://www.e-unwto.org/doi/abs/10.5555/unwtotfb0000270020142018202001); and 5) bachelor’s or equivalent (ISCED [International Standard Classification of Education] 6) (%) in 2016–2018 from the UNESCO [United Nations Educational, Scientific and Cultural Organization] (http://uis.unesco.org/en/topic/educational-attainment) (**Table 1**). Restricted maximum-likelihood meta-regression in the random-effects model was performed using Comprehensive Meta-Analysis version 3 (Biostat, Englewood, NJ, USA). A meta-regression graph depicted the COVID-19 prevalence (%) (plotted as the logarithm-transformed prevalence on the y-axis) as a function of a given covariate (plotted on the x-axis). To adjust for other covariates, we conducted the hierarchical multivariate models in addition to the univariate model.

**Table 1.** Data from Worldwide Nations.

| **Nation** | **Hypertension prevalence (%)** | **Obesity prevalence (%)** | **Diabetes prevalence (%)** | **Annual PM2.5 concentrations in urban areas (µg/m³)** | **Daily ambient ultraviolet radiation (J/m²)** | **Population using safely-managed drinking-water services (%)** | **Population using safely-managed sanitation services (%)** | **Population using hand-washing facility with soap/water (%)** | **Inbound tourism (thousands)** | **Bachelor's or equivalent (ISCED 6) (%)** |
| --- | --- | --- | --- | --- | --- | --- | --- | --- | --- | --- |
| Afghanistan | 30.6 | 5.5 | 9.2 | 59.900 | 4132 | N/A | N/A | 38 | N/A | N/A |
| Albania | 29.0 | 21.7 | 9.0 | 18.200 | 2542 | 70 | 40 | N/A | 5927 | N/A |
| Algeria | 25.1 | 27.4 | 6.7 | 34.500 | 3253 | N/A | 18 | 84 | 2657 | N/A |
| Andorra | 18.7 | 25.6 | 7.7 | 11.500 | 2469 | 91 | 100 | N/A | 8328 | 10.2 |
| Angola | 29.7 | 8.2 | 4.5 | 28.400 | 5287 | N/A | N/A | 27 | N/A | N/A |
| Antigua and Barbuda | 23.4 | 18.9 | 13.1 | 18.000 | 5148 | N/A | N/A | N/A | 1062 | N/A |
| Argentina | 22.6 | 28.3 | 5.9 | 11.700 | 3476 | N/A | N/A | N/A | 10394 | 20.0 |
| Armenia | 25.5 | 20.2 | 6.1 | 32.900 | 2899 | 86 | 48 | 94 | N/A | 24.1 |
| Australia | 15.2 | 29.0 | 5.6 | 7.300 | 3206 | N/A | 76 | N/A | 9246 | 23.3 |
| Austria | 21.0 | 20.1 | 6.6 | 13.100 | 1888 | 99 | 97 | N/A | N/A | 2.8 |
| Azerbaijan | 24.5 | 19.9 | 6.1 | 18.500 | 2702 | 74 | N/A | 83 | 2850 | 15.7 |
| Bahamas | 20.9 | 31.6 | 8.8 | 19.000 | 4119 | N/A | N/A | N/A | 6622 | N/A |
| Bahrain | 21.4 | 29.8 | 15.6 | 69.000 | 4419 | 99 | 96 | N/A | 12045 | 20.2 |
| Bangladesh | 24.7 | 3.6 | 9.2 | 58.600 | 4029 | 55 | N/A | 35 | N/A | 5.3 |
| Barbados | 24.4 | 23.1 | 13.4 | 22.400 | 5787 | N/A | N/A | N/A | 1356 | N/A |
| Belarus | 27.1 | 24.5 | 5.0 | 19.300 | 1795 | 95 | 81 | N/A | 11502 | N/A |
| Belgium | 17.5 | 22.1 | 4.6 | 13.000 | 1645 | 100 | 97 | N/A | N/A | 19.8 |
| Belize | 22.7 | 24.1 | 17.1 | 20.900 | 4639 | N/A | N/A | 90 | 1697 | N/A |
| Benin | 27.7 | 9.6 | 1.0 | 30.400 | 5168 | N/A | N/A | 11 | 322 | N/A |
| Bhutan | 28.1 | 6.4 | 10.3 | 35.400 | 4180 | 36 | N/A | N/A | N/A | 7.9 |
| Bolivia (Plurinational State of) | 17.9 | 20.2 | 6.8 | 23.300 | 5344 | N/A | 23 | 25 | N/A | N/A |
| Bosnia and Herzegovina | 30.8 | 17.9 | 9.0 | 29.700 | 2205 | 89 | 22 | N/A | N/A | 6.6 |
| Botswana | 29.6 | 18.9 | 5.8 | 20.900 | 4868 | N/A | N/A | N/A | 1775 | N/A |
| Brazil | 23.3 | 22.1 | 10.4 | 11.800 | 4552 | N/A | 49 | N/A | N/A | 15.6 |
| Brunei Darussalam | 18.9 | 14.1 | 13.3 | 5.800 | 5148 | N/A | N/A | N/A | 4521 | N/A |
| Bulgaria | 28.4 | 25.0 | 6.0 | 20.800 | 2331 | 97 | 64 | N/A | 12368 | 24.7 |
| Burkina Faso | 32.6 | 5.6 | 7.3 | 36.300 | 5567 | N/A | N/A | 12 | N/A | N/A |
| Burundi | 29.2 | 5.4 | 5.1 | 35.600 | 5111 | N/A | N/A | 6 | N/A | 0.9 |
| Cabo Verde | 29.5 | 11.8 | 2.4 | 31.600 | 5372 | N/A | N/A | N/A | N/A | N/A |
| Cambodia | 26.1 | 3.9 | 6.4 | 24.900 | 5152 | 26 | N/A | 66 | N/A | N/A |
| Cameroon | 24.8 | 11.4 | 6.0 | 65.400 | 5185 | N/A | N/A | 9 | N/A | N/A |
| Canada | 13.2 | 29.4 | 7.6 | 6.700 | 1887 | 99 | 82 | N/A | 31274 | 18.7 |
| Central African Republic | 31.2 | 7.5 | 6.0 | 51.200 | 5498 | N/A | N/A | N/A | N/A | N/A |
| Chad | 32.9 | 6.1 | 6.0 | 50.800 | 5669 | N/A | N/A | 6 | N/A | N/A |
| Chile | 20.9 | 28.0 | 8.6 | 23.100 | 3982 | 99 | 77 | N/A | 6603 | 13.1 |
| China | 19.2 | 6.2 | 9.2 | 51.000 | 2908 | N/A | 72 | N/A | 158606 | N/A |
| Colombia | 19.2 | 22.3 | 7.4 | 17.200 | 5385 | 73 | 17 | 65 | 4282 | 8.2 |
| Comoros | 27.9 | 7.8 | 12.3 | 18.600 | 5524 | N/A | N/A | N/A | N/A | N/A |
| Congo | 26.2 | 9.6 | 6.0 | 36.400 | 4943 | 45 | N/A | 48 | 158 | N/A |
| Costa Rica | 18.7 | 25.7 | 9.1 | 16.700 | 4884 | 94 | N/A | N/A | 3300 | 18.4 |
| Côte d'Ivoire | 27.2 | 10.3 | 2.4 | 23.900 | 4931 | 37 | N/A | 19 | 1965 | N/A |
| Croatia | 32.4 | 24.4 | 5.4 | 17.600 | 1976 | 90 | 58 | N/A | 57668 | N/A |
| Cuba | 19.0 | 24.6 | 9.6 | 21.600 | 4401 | N/A | 44 | 85 | 4712 | N/A |
| Cyprus | 19.8 | 21.8 | 9.0 | 17.100 | 3439 | 100 | 75 | N/A | 4024 | 16.4 |
| Czechia | 27.9 | 26.0 | 7.0 | 15.600 | 1707 | 98 | 94 | N/A | 34701 | 4.6 |
| Democratic Republic of the Congo | 28.5 | 6.7 | 6.0 | 37.400 | 5100 | N/A | N/A | 4 | N/A | 3.3 |
| Denmark | 20.6 | 19.7 | 8.3 | 10.300 | 1691 | 97 | 95 | N/A | 30801 | 19.0 |
| Djibouti | 26.8 | 13.5 | 5.1 | 41.000 | 5461 | N/A | 36 | N/A | N/A | N/A |
| Dominica | 22.5 | 27.9 | 11.6 | 18.800 | 5464 | N/A | N/A | N/A | 198 | N/A |
| Dominican Republic | 21.5 | 27.6 | 8.6 | 13.300 | 4880 | N/A | N/A | 55 | 7576 | 16.9 |
| Ecuador | 17.9 | 19.9 | 5.5 | 15.500 | 4929 | 75 | 42 | 81 | 2535 | 10.9 |
| Egypt | 25.0 | 32.0 | 17.2 | 79.600 | 4202 | N/A | 61 | 90 | 11346 | 5.5 |
| El Salvador | 18.7 | 24.6 | 8.8 | 23.800 | 5364 | N/A | N/A | 91 | 2536 | 6.7 |
| Equatorial Guinea | 28.4 | 8.0 | 6.0 | 49.100 | 4635 | N/A | N/A | N/A | N/A | N/A |
| Eritrea | 29.1 | 5.0 | 5.1 | 41.100 | 5914 | N/A | N/A | N/A | 142 | N/A |
| Estonia | 27.4 | 21.2 | 4.2 | 7.000 | 1781 | 93 | 97 | N/A | 6071 | 3.3 |
| Eswatini | 29.8 | 16.5 | 4.5 | 16.200 | 3900 | N/A | N/A | 24 | 1277 | N/A |
| Ethiopia | 30.3 | 4.5 | 4.3 | 34.000 | 5929 | 11 | N/A | 8 | N/A | N/A |
| Fiji | 21.7 | 30.2 | 14.7 | 10.500 | 4431 | N/A | N/A | N/A | 1058 | 3.7 |
| Finland | 19.4 | 22.2 | 5.6 | 6.500 | 1494 | 100 | 100 | N/A | N/A | 12.4 |
| France | 22.0 | 21.6 | 4.8 | 12.400 | 1907 | 98 | 88 | N/A | 211998 | 8.5 |
| Gabon | 25.5 | 15.0 | 6.0 | 37.800 | 4654 | N/A | N/A | N/A | N/A | N/A |
| Gambia | 29.1 | 10.3 | 1.9 | 32.300 | 5358 | N/A | N/A | 8 | N/A | N/A |
| Georgia | 26.3 | 21.7 | 5.8 | 24.000 | 2489 | 80 | 27 | N/A | 7203 | 6.9 |
| Germany | 19.9 | 22.3 | 10.4 | 11.900 | 1812 | 100 | 97 | N/A | N/A | 13.9 |
| Ghana | 23.7 | 10.9 | 2.5 | 31.100 | 5166 | 36 | N/A | 41 | N/A | N/A |
| Greece | 19.1 | 24.9 | 4.7 | 16.400 | 2753 | 100 | 90 | N/A | 33072 | 17.7 |
| Grenada | 24.3 | 21.3 | 10.7 | 21.800 | 4875 | 87 | N/A | N/A | 528 | N/A |
| Guatemala | 21.2 | 21.2 | 10.0 | 24.200 | 5141 | 56 | N/A | 77 | 2406 | N/A |
| Guinea | 30.3 | 7.7 | 2.4 | 22.200 | 5391 | N/A | N/A | 17 | N/A | N/A |
| Guinea-Bissau | 30.3 | 9.5 | 2.4 | 26.500 | 5319 | N/A | N/A | 6 | N/A | N/A |
| Guyana | 23.1 | 20.2 | 11.6 | 21.600 | 5203 | N/A | N/A | 77 | N/A | N/A |
| Haiti | 24.5 | 22.7 | 6.7 | 14.700 | 5016 | N/A | N/A | 23 | 1333 | N/A |
| Honduras | 21.4 | 21.4 | 7.3 | 21.500 | 4924 | N/A | N/A | N/A | 2303 | 9.5 |
| Hungary | 30.0 | 26.4 | 6.9 | 16.300 | 1932 | 90 | 96 | N/A | 57667 | 11.8 |
| Iceland | 19.7 | 21.9 | 5.8 | 5.900 | 957 | 100 | 82 | N/A | 2488 | N/A |
| India | 25.8 | 3.9 | 10.4 | 68.000 | 4514 | N/A | N/A | 60 | N/A | N/A |
| Indonesia | 23.8 | 6.9 | 6.3 | 16.400 | 5220 | N/A | N/A | 64 | 15810 | 8.8 |
| Iran (Islamic Republic of) | 19.7 | 25.8 | 9.6 | 34.400 | 4038 | 92 | N/A | N/A | 7295 | 13.7 |
| Iraq | 25.2 | 30.4 | 8.8 | 60.100 | 3825 | 59 | 41 | 95 | N/A | N/A |
| Ireland | 19.7 | 25.3 | 3.2 | 8.700 | 1509 | 97 | 82 | N/A | N/A | 18.6 |
| Israel | 16.6 | 26.1 | 9.7 | 19.400 | 3682 | 100 | 94 | N/A | 4389 | N/A |
| Italy | 21.2 | 19.9 | 5.0 | 15.700 | 2444 | 95 | 96 | N/A | 93228.6 | N/A |
| Jamaica | 21.8 | 24.7 | 11.3 | 13.600 | 4942 | N/A | N/A | N/A | 4319 | N/A |
| Japan | 17.6 | 4.3 | 5.6 | 11.800 | 2521 | 98 | 99 | N/A | 31192 | N/A |
| Jordan | 21.0 | 35.5 | 12.7 | 31.700 | 4026 | 94 | 81 | N/A | 4922 | N/A |
| Kazakhstan | 27.1 | 21.0 | 6.1 | 14.500 | 2257 | 90 | N/A | 99 | 8789 | 34.0 |
| Kenya | 26.7 | 7.1 | 3.1 | 25.800 | 5803 | N/A | N/A | 25 | 1449 | N/A |
| Kuwait | 23.6 | 37.9 | 12.2 | 58.900 | 4214 | 100 | 100 | N/A | 8508 | 9.8 |
| Kyrgyzstan | 26.7 | 16.6 | 6.1 | 17.400 | 3094 | 68 | N/A | 89 | 6947 | N/A |
| Lao People's Democratic Republic | 24.8 | 5.3 | 6.4 | 25.500 | 4735 | 16 | 58 | 50 | 4186 | N/A |
| Latvia | 29.4 | 23.6 | 5.0 | 14.400 | 1671 | 95 | 86 | N/A | 7775 | 15.4 |
| Lebanon | 20.7 | 32.0 | 11.2 | 30.700 | 2953 | 48 | 22 | N/A | N/A | N/A |
| Lesotho | 29.0 | 16.6 | 4.5 | 28.100 | 4439 | N/A | N/A | 2 | 1173 | N/A |
| Liberia | 28.3 | 9.9 | 2.4 | 17.000 | 4926 | N/A | N/A | 1 | N/A | N/A |
| Libya | 23.7 | 32.5 | 10.2 | 41.700 | 3861 | N/A | 26 | N/A | N/A | N/A |
| Lithuania | 29.3 | 26.3 | 3.8 | 12.300 | 1801 | 92 | 91 | N/A | 6115 | 21.2 |
| Luxembourg | 21.9 | 22.6 | 5.0 | 10.400 | 1687 | 100 | 97 | N/A | N/A | N/A |
| Madagascar | 28.1 | 5.3 | 4.5 | 22.500 | 4771 | N/A | N/A | N/A | N/A | N/A |
| Malawi | 28.9 | 5.8 | 4.5 | 21.900 | 5019 | N/A | N/A | 9 | N/A | N/A |
| Malaysia | 22.9 | 15.6 | 16.7 | 17.300 | 5225 | 93 | 89 | N/A | N/A | 9.7 |
| Maldives | 24.4 | 8.6 | 9.2 | 7.700 | 10224 | N/A | N/A | 96 | N/A | N/A |
| Mali | 32.6 | 8.6 | 2.4 | 29.000 | 5617 | N/A | N/A | 52 | N/A | 2.1 |
| Malta | 19.4 | 28.9 | 8.3 | 14.000 | 3091 | 100 | 93 | N/A | 3232 | 11.8 |
| Mauritania | 31.7 | 12.7 | 7.1 | 41.700 | 5547 | N/A | N/A | 43 | 1002 | N/A |
| Mauritius | 25.0 | 10.8 | 22.0 | 13.500 | 5055 | N/A | N/A | N/A | 1431 | 7.6 |
| Mexico | 19.7 | 28.9 | 13.5 | 20.900 | 4974 | 43 | 50 | 88 | 96497 | 14.5 |
| Monaco | N/A | N/A | 2.9 | 12.200 | 1907 | 100 | 100 | N/A | N/A | N/A |
| Mongolia | 29.0 | 20.6 | 4.7 | 49.500 | 2226 | 24 | N/A | 71 | 598 | N/A |
| Montenegro | 29.1 | 23.3 | 9.0 | 19.300 | N/A | 94 | N/A | N/A | N/A | N/A |
| Morocco | 26.1 | 26.1 | 7.0 | 31.100 | 3568 | 70 | 39 | N/A | 12489 | N/A |
| Mozambique | 29.1 | 7.2 | 3.3 | 18.400 | 4646 | N/A | N/A | N/A | 2870 | 1.6 |
| Myanmar | 24.6 | 5.8 | 3.9 | 34.600 | 4565 | N/A | N/A | 79 | N/A | N/A |
| Namibia | 28.5 | 17.2 | 4.5 | 21.000 | 5305 | N/A | N/A | 45 | 1581 | N/A |
| Nepal | 29.4 | 4.1 | 7.2 | 99.500 | 4130 | 27 | N/A | 48 | N/A | N/A |
| Netherlands | 18.7 | 20.4 | 5.4 | 12.100 | 1662 | 100 | 97 | N/A | N/A | 19.1 |
| New Zealand | 16.2 | 30.8 | 6.2 | 5.800 | 2487 | 100 | 89 | N/A | 3858 | 22.5 |
| Nicaragua | 20.8 | 23.7 | 11.4 | 19.000 | 5078 | 52 | N/A | N/A | 1412 | N/A |
| Niger | 33.4 | 5.5 | 2.4 | 73.000 | 5811 | N/A | 10 | N/A | N/A | N/A |
| Nigeria | 23.9 | 8.9 | 3.1 | 46.300 | 5251 | 20 | 27 | 42 | 5265 | N/A |
| North Macedonia | 28.5 | 22.4 | 9.3 | 33.000 | N/A | 81 | 17 | N/A | N/A | N/A |
| Norway | 19.7 | 23.1 | 5.3 | 7.800 | 1439 | 98 | 76 | N/A | N/A | 16.4 |
| Oman | 24.8 | 27.0 | 10.1 | 36.200 | 5102 | 90 | N/A | 97 | 3242 | N/A |
| Pakistan | 30.5 | 8.6 | 19.9 | 56.200 | 4227 | 35 | N/A | 60 | N/A | 6.8 |
| Panama | 19.9 | 22.7 | 7.7 | 12.000 | 4898 | v | N/A | N/A | 2487 | N/A |
| Papua New Guinea | 25.6 | 21.3 | 17.9 | 11.500 | 5377 | N/A | N/A | N/A | 195 | N/A |
| Paraguay | 24.6 | 20.3 | 9.6 | 11.700 | 4038 | 64 | 58 | 80 | 4183 | 6.1 |
| Peru | 13.7 | 19.7 | 6.6 | 29.000 | 5906 | 50 | 43 | N/A | 5384 | 10.0 |
| Philippines | 22.6 | 6.4 | 7.1 | 18.700 | 4928 | 47 | 52 | 78 | N/A | 15.6 |
| Poland | 28.7 | 23.1 | 6.1 | 21.500 | 1749 | 100 | 93 | N/A | 85946 | 5.7 |
| Portugal | 24.4 | 20.8 | 9.8 | 8.100 | 2585 | 95 | 85 | N/A | N/A | 5.0 |
| Qatar | 22.4 | 35.1 | 15.6 | 91.700 | 4905 | 96 | 96 | N/A | N/A | 17.6 |
| Republic of Korea | 11.0 | 4.7 | 6.9 | 24.700 | 2535 | 98 | 100 | N/A | 15347 | N/A |
| Republic of Moldova | 29.8 | 18.9 | 5.7 | 16.500 | 1910 | 73 | N/A | N/A | N/A | 19.0 |
| Romania | 30.0 | 22.5 | 6.9 | 15.400 | 2403 | 82 | 77 | N/A | 11720 | N/A |
| Russian Federation | 27.2 | 23.1 | 6.1 | 14.700 | 2071 | 76 | 61 | N/A | 24551 | N/A |
| Rwanda | 26.7 | 5.8 | 5.1 | 40.700 | 1795 | N/A | N/A | 5 | 1307 | 3.8 |
| Saint Kitts and Nevis | 25.3 | 22.9 | 13.3 | 12.300 | 5130 | N/A | N/A | N/A | 1278 | N/A |
| Saint Lucia | 27.1 | 19.7 | 11.6 | 21.200 | 5009 | N/A | N/A | N/A | 1165 | N/A |
| Saint Vincent and the Grenadines | 23.3 | 23.7 | 11.6 | 21.400 | 5164 | N/A | N/A | N/A | 356 | N/A |
| San Marino | N/A | N/A | 5.9 | 13.400 | 4966 | 100 | 77 | N/A | 1874 | 4.3 |
| São Tomé and Príncipe | 25.8 | 12.4 | 2.4 | 25.200 | 1932 | N/A | N/A | 41 | N/A | N/A |
| Saudi Arabia | 23.3 | 35.4 | 15.8 | 86.700 | 4422 | N/A | 78 | N/A | 17570 | 23.5 |
| Senegal | 30.2 | 8.8 | 2.4 | 39.700 | 5384 | N/A | 21 | 24 | 1376 | 1.2 |
| Serbia | 29.5 | 21.5 | 9.0 | 24.700 | N/A | 75 | 25 | N/A | N/A | 12.7 |
| Seychelles | 23.5 | 14.0 | 12.3 | 18.600 | 5356 | N/A | N/A | N/A | 405 | N/A |
| Sierra Leone | 30.3 | 8.7 | 2.4 | 20.600 | 5570 | 10 | 13 | 19 | 66 | N/A |
| Singapore | 14.6 | 6.1 | 2.4 | 18.300 | 5087 | 100 | 100 | N/A | 18508 | 31.6 |
| Slovakia | 28.5 | 20.5 | 6.5 | 18.000 | 3979 | 100 | 83 | N/A | 23092 | 2.1 |
| Slovenia | 30.5 | 20.2 | 5.9 | 16.400 | 1795 | 98 | 83 | N/A | N/A | 5.4 |
| Somalia | 32.9 | 8.3 | 5.1 | 28.000 | 4071 | N/A | N/A | 10 | N/A | N/A |
| South Africa | 26.9 | 28.3 | 12.7 | 24.300 | 5773 | N/A | N/A | 44 | 15004 | 6.8 |
| South Sudan | N/A | N/A | 10.2 | 40.900 | 4111 | N/A | N/A | N/A | N/A | N/A |
| Spain | 19.2 | 23.8 | 6.9 | 9.800 | 2705 | 98 | 97 | N/A | 124060 | 9.0 |
| Sri Lanka | 22.4 | 5.2 | 10.7 | 15.100 | 5264 | N/A | N/A | N/A | 2521 | 3.6 |
| Sudan | N/A | N/A | 22.1 | 46.800 | 5783 | N/A | N/A | 23 | 3093 | N/A |
| Suriname | 22.4 | 26.4 | 12.5 | 25.800 | 4799 | N/A | N/A | N/A | 279 | N/A |
| Sweden | 19.3 | 20.6 | 4.8 | 6.100 | 1587 | 100 | 93 | N/A | N/A | 12.4 |
| Switzerland | 18.0 | 19.5 | 5.7 | 10.400 | 2158 | 95 | 100 | N/A | N/A | 18.8 |
| Syrian Arab Republic | 24.5 | 27.8 | 13.5 | 37.400 | 3501 | N/A | N/A | 71 | N/A | N/A |
| Tajikistan | 26.1 | 14.2 | 6.1 | 42.800 | 3538 | 48 | N/A | 73 | 1035 | 0.9 |
| Thailand | 22.3 | 10.0 | 7.0 | 26.600 | 4862 | N/A | N/A | 84 | N/A | 12.8 |
| The United Kingdom | 15.2 | 27.8 | 3.9 | 10.600 | 1576 | 100 | 98 | N/A | 37905 | 17.2 |
| Timor-Leste | 27.6 | 3.8 | 6.7 | 18.200 | 5637 | N/A | N/A | 28 | N/A | N/A |
| Togo | 28.9 | 8.4 | 2.4 | 31.200 | 5307 | N/A | N/A | 10 | N/A | N/A |
| Trinidad and Tobago | 25.8 | 18.6 | 11.0 | 22.400 | 5245 | N/A | N/A | N/A | 501 | N/A |
| Tunisia | 23.2 | 26.9 | 8.5 | 35.700 | 3262 | 93 | 78 | 79 | N/A | 15.2 |
| Turkey | 20.3 | 32.1 | 11.1 | 41.200 | 2924 | N/A | 65 | N/A | 46113 | 16.7 |
| Uganda | 27.3 | 5.3 | 2.5 | 48.700 | 5499 | 7 | N/A | 21 | N/A | N/A |
| Ukraine | 27.1 | 24.1 | 6.1 | 19.400 | 1843 | 92 | 68 | N/A | 14342 | N/A |
| United Arab Emirates | 21.1 | 31.7 | 16.3 | 37.200 | 4862 | N/A | 96 | N/A | N/A | 37.2 |
| United Republic of Tanzania | 27.3 | 8.4 | 5.7 | 25.100 | 5483 | N/A | 25 | 48 | 1506 | N/A |
| United States of America | 12.9 | 36.2 | 10.8 | 7.600 | 2736 | 100 | 90 | N/A | 169324.918 | 21.9 |
| Uruguay | 20.7 | 27.9 | 7.3 | 8.700 | 3235 | N/A | N/A | N/A | 3954 | 9.4 |
| Uzbekistan | 25.6 | 16.6 | 6.5 | 28.900 | 3172 | 59 | N/A | N/A | 5346 | 16.3 |
| Venezuela (Bolivarian Republic of) | 18.6 | 25.6 | 7.0 | 16.800 | 5211 | N/A | 24 | N/A | 429 | 24.1 |
| Viet Nam | 23.4 | 2.1 | 6.0 | 30.100 | 4293 | N/A | N/A | 86 | 15498 | N/A |
| Yemen | 30.7 | 17.1 | 5.4 | 44.300 | 6089 | N/A | N/A | 50 | N/A | N/A |
| Zambia | 27.1 | 8.1 | 4.5 | 23.800 | 5265 | N/A | N/A | 14 | N/A | N/A |
| Zimbabwe | 28.2 | 15.5 | 1.8 | 19.100 | 4918 | N/A | N/A | 37 | 2580 | 2.6 |

Abbreviation: GDP, Gross Domestic Product; GNI, Gross National Income; ISCED, International Standard Classification of Education; PM2.5, particulate matter 2.5; PPP, Purchasing Power Parity.

## Results

Results of the meta-regression were summarized in **Table 2**. A slope (coefficient) of the meta-regression line for the COVID-19 prevalence in the multivariable models was significantly negative for population ages 0-14 (coefficient, –0.0636; *P* = .0021; **Figure 1A**) and positive for obesity prevalence (coefficient, 0.0411; *P* = .0099; **Figure 1B**) and annual PM2.5 concentrations in urban areas (coefficient, 0.0158; *P* = .0454; **Figure 1C**), which would indicate that the COVID-19 prevalence decreases significantly as children increase and that the COVID-19 prevalence increases significantly as the obese and PM2.5 increase.

**Table 2.**
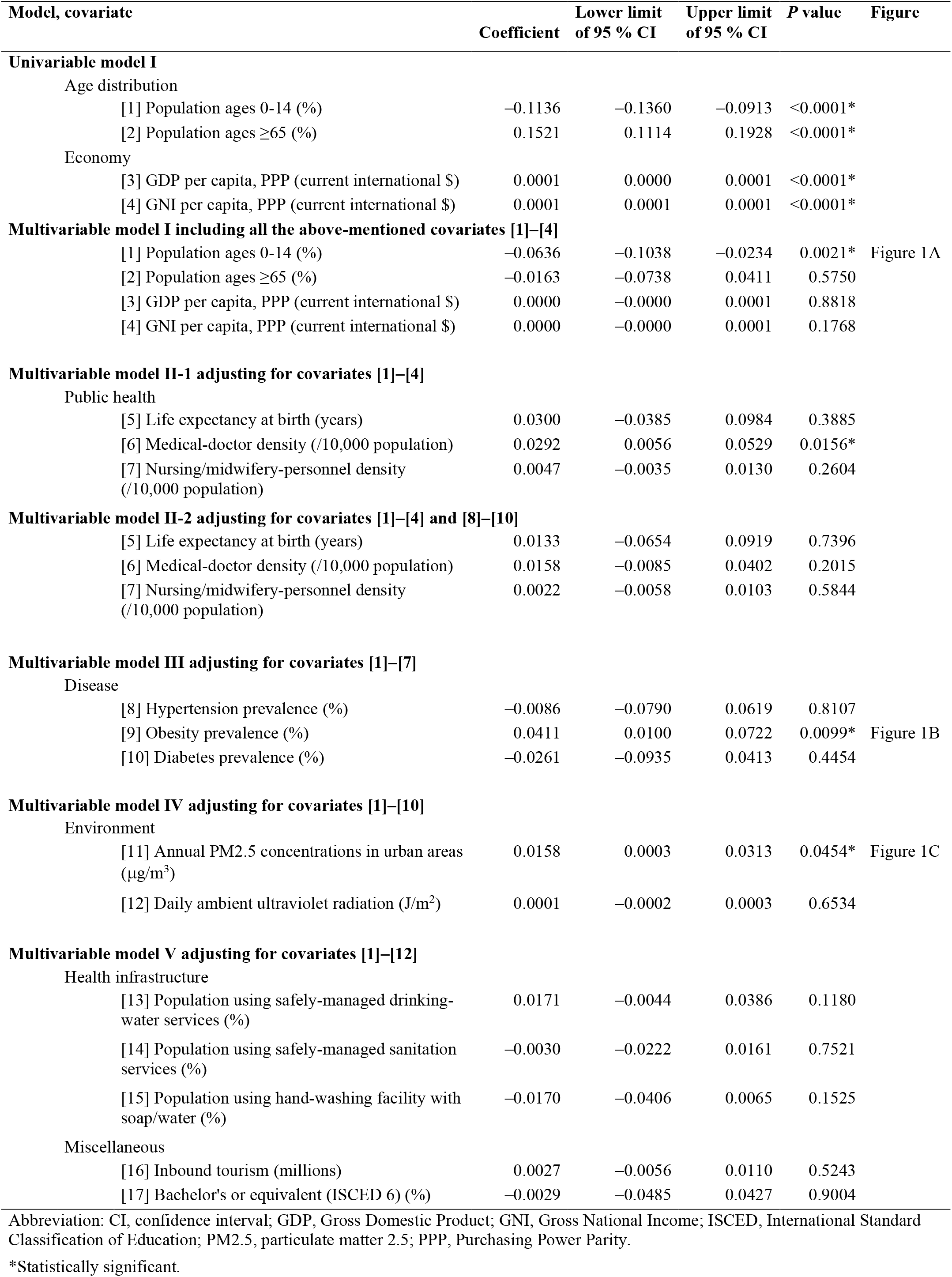
Meta-regression Summary.

**Figure 1.**
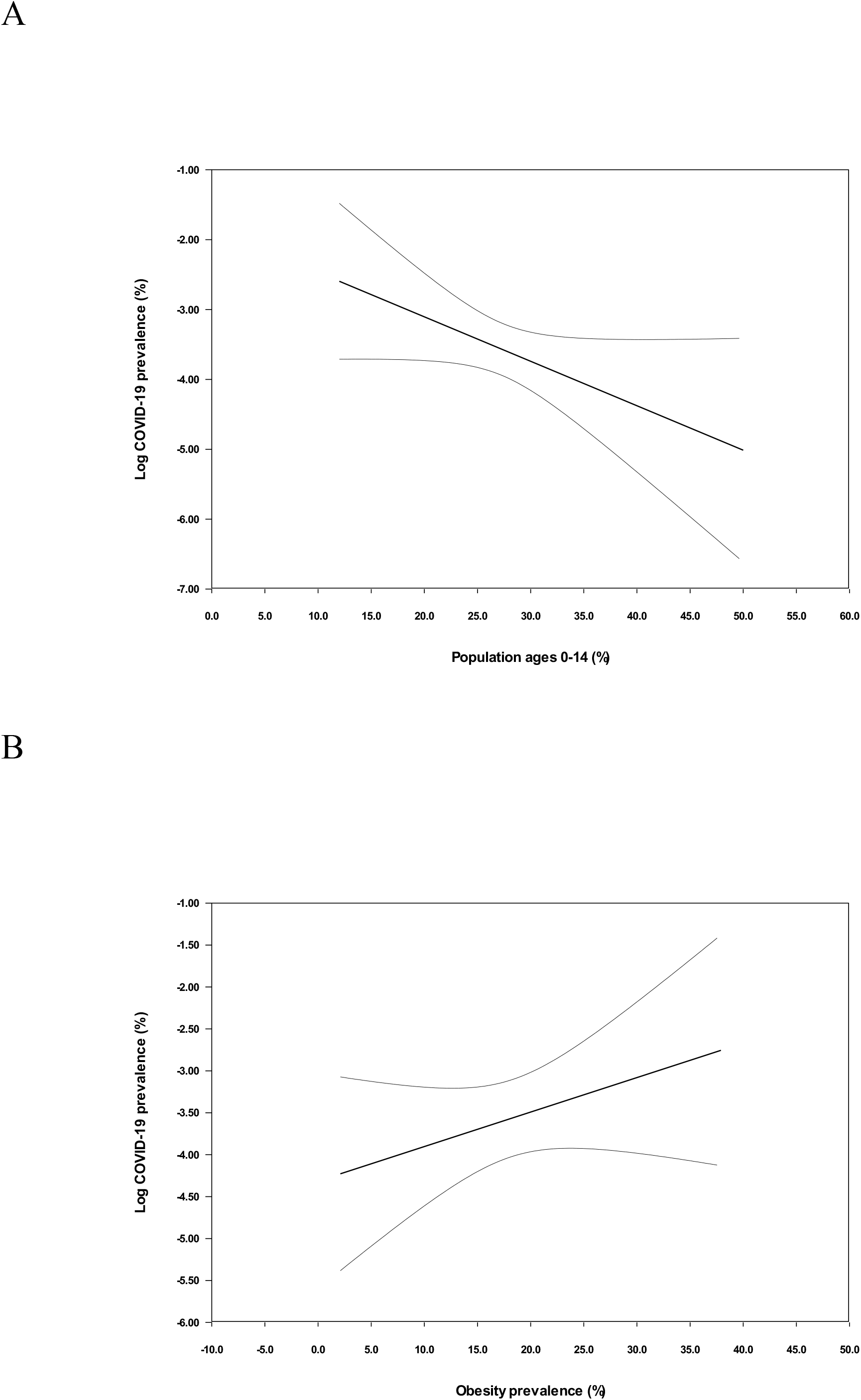
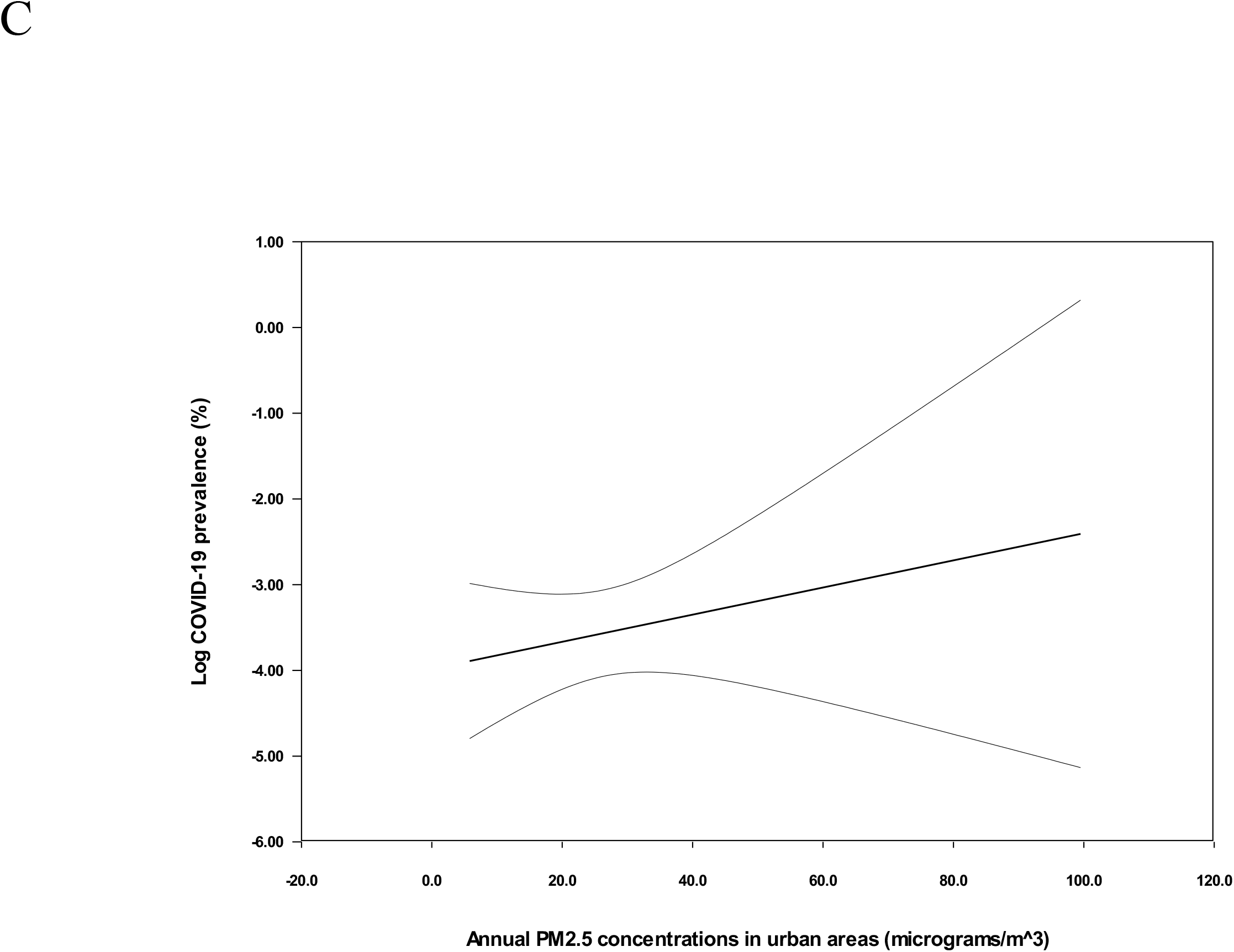
Meta-Regression Lines with their 95% Confidence-Interval Curves Depicting the COVID-19 Prevalence (Plotted as the Logarithm-Transformed Prevalence on the Y-Axis) as a Function of a Given Covariate (Plotted on the X-Axis)

## Discussion

The present meta-regression to screen potential risk and protective factors suggests that children may be negatively and independently, and obesity/PM2.5 may be positively and independently associated with SARS-CoV-2 infection. Our findings could be strengthened by low case fatality in children with COVID-19 (only one death in a total of 1124 cases)^3^ and obesity predicting poor prognosis of COVID-19^4^ demonstrated in recent systematic reviews. The present results, however, never denote directly that, for instance, the obese are at high risk for SARS-CoV-2 infection, which should be noted. Our findings demonstrate simply that the COVID-19 prevalence is higher in the nation where the obese are more. To determine whether, for instance, the obese are at high risk for SARS-CoV-2 infection, two approaches are considered. First, potential risk and protective factors could be investigated in both COVID-19 patients and non-COVID-19 subjects in a community. It is never facile, however, to investigate the factors in the non-COVID-19 (healthy) subjects in the community. Second, the factors could be examined in both COVID-19 patients and control subjects. In this occasion, however, the control subjects, who should be well comparable to the COVID-19 patients, must be strictly selected. Meta-regression using “weighted” data from multiple clinical trials differs from simple regression using individual-patient data from a single study and accordingly can generate an equation of a “best-fit” regression line to express the relation between an outcome and a covariate.^5^ Meta-regression applied in the present study may be alternative to the above-mentioned approaches and could be of use at least to screen the factors.

## Conclusion

Children (negatively) and obesity/PM2.5 (positively) may be independently associated with SARS-CoV-2 infection.

## Data Availability

The datasets generated during and/or analysed during the current study are available from the corresponding author on reasonable request.

## References

1. Liu H, Chen S, Liu M, Nie H, Lu H. Comorbid Chronic Diseases are Strongly Correlated with Disease Severity among COVID-19 Patients: A Systematic Review and Meta-Analysis. Aging Dis. 2020 May 9;11(3):668–678. doi: 10.14336/AD.2020.0502

2. Deeks JJ, Higgins JPT, Altman DG, eds. Chapter 10: Analysing data and undertaking meta-analyses. In: Higgins JPT, Thomas J, Chandler J, et al, eds. Cochrane Handbook for Systematic Reviews of Interventions version 6.0 (updated July 2019). Cochrane, 2019. https://training.cochrane.org/handbook. Accessed June 1, 2020.

3. de Souza TH, Nadal JA, Nogueira RJN, Pereira RM, Brandão MB. Clinical Manifestations of Children with COVID-19: a Systematic Review. Pediatr Pulmonol. Published online Jun 3, 2020. doi:10.1002/ppul.24885

4. Tamara A, Tahapary DL. Obesity as a predictor for a poor prognosis of COVID-19: A systematic review. Diabetes Metab Syndr. 2020;14(4):655–659. doi:10.1016/j.dsx.2020.05.020

5. Baker WL, White CM, Cappelleri JC, Kluger J, Coleman CI; Health Outcomes, Policy, and Economics (HOPE) Collaborative Group. Understanding heterogeneity in meta-analysis: the role of meta-regression. Int J Clin Pract. 2009;63(10):1426–1434. doi:10.1111/j.1742-1241.2009.02168.x

